# Beyond the Dose: Introducing LOGOS for Dosage Regimen Design in Therapeutics

**DOI:** 10.64898/2026.10.17.26365797

**Authors:** Vangelis D. Karalis

**Affiliations:** Department of Pharmacy, School of Health Sciences, National and Kapodistrian University of Athens, 15784 Athens, Greece; Institute of Applied and Computational Mathematics, Foundations for Research and Technology Hellas (FORTH), 70013, Heraklion, Greece

**Keywords:** regimen design, pharmacokinetics, pharmacodynamics, optimal control, treatment cessation, recovery

## Abstract

The aim of this study was to develop a framework for assessing the incremental value of decisions on drug amount, timing, input structure, cessation and restart. The LOGOS (Longitudinal Optimization of Global Outcomes and Strategies) approach combines matched nested regimen classes, mechanism-specific benchmarks and evidence requirements in a single longitudinal comparison. Thus, the contribution of additional regimen freedom is assessed separately from improvements due to different resources, constraints or evaluation horizons. Nested feasible classes have nondecreasing objective suprema, whereas a strict gain requires a feasible witness above a valid bound for the smaller class. The approach is illustrated through analytical examples of periodic smoothing, matched desensitization, relaxed exposure allocation, finite-course loading and safety after cessation. In a normalized finite Emax model, integrated benefit increased from 0.629606 for three equal boluses to 0.672945 after optimization of amounts, 0.677043 for an amount-and-time candidate and 0.679553 for a globally certified loading-maintenance-coast solution. The adjacent observed gains were 6.88%, 0.61% and 0.37%, respectively, and all schedules satisfied the full-tail injury bound. In addition, a periodic factorial example separated response curvature from a joint recovery and repair process, including a null case and results dependent on feasibility constraints. The LOGOS framework allows assessment of whether additional regimen flexibility improves benefit, provides no added value or restores feasibility. These dimensionless examples support conditional mathematical and model-based conclusions; clinical superiority and safe restart in patients have not been established.

## 1 Introduction

Drug administration gives rise to a time-dependent trajectory through a biological system. The consequence of a dose depends on residual exposure, current responsiveness, accumulated injury, disease state and the time available for benefit or recovery. A regimen therefore defines not only an amount of drug but also when and how drug enters the system and how long treatment lasts, when it ends and under what conditions it will be resumed. The question is which attainable trajectory best achieves the specified therapeutic goal.

Model-based dosing and optimal control already provide substantial support for this question. OptiDose treats individualized PK/PD dosing as an optimal control problem [1]. The improved method addresses efficacy and safety targets including state constraints and dosing-interval design [2]. Disease-specific optimization has also addressed treatment switching and drug holidays with toxicity constraints, such as chronic myeloid leukemia which is treated with imatinib, dasatinib and nilotinib [3]. Optimization, toxicity modeling and adjustment of different dosing variables cannot, by themselves, establish the contribution of a new framework.

LOGOS (Longitudinal Optimization of Global Outcomes and Strategies) provides an integrated protocol for assessing the value of additional regimen freedom. It combines nested admissible classes under matched objectives, resource rules and constraints with mechanism-specific positive and null benchmarks. It also distinguishes feasible candidates from class-level bounds and global certificates. For each proposed freedom, the comparison first establishes whether this choice is already available in the reference regimen. Its effect on benefit or feasibility is then assessed, together with the underlying mechanism and the attainability and safety of the trajectory over the declared horizon. In this way, the complexity of whole-regimen design is related to the evidence required to support it. It should be stated that a favorable static exposure distribution does not necessarily correspond to an achievable schedule. Similarly, an administration pause does not establish biological recovery, and a better numerical candidate does not establish a global optimum.

The term longitudinal describes the evolution of the treatment system over the evaluation and safety horizons. Global outcomes refer to the combined therapeutic consequences selected for assessment, without implying global optimality of every numerical solution. Strategies include both predetermined schedules and policies in which decisions are adjusted according to information available during treatment. The underlying pharmacokinetic and pharmacodynamic description of the system is denoted by PK/PD.

Several established optimization approaches already allow adjustment of multiple dosing variables [1–3]. In the present analysis, the question is whether an additional freedom provides value when the objective, model, resource rule and relevant constraints are kept the same. Bayesian model-informed precision dosing supports individualized prediction and adjustment [7,12,14]. Additionally, adaptive control and trial designs address decisions based on observations and the selection of doses and schedules [8,9,13]. The LOGOS approach allows these strategies to be assessed within a common comparison, without considering their labels as evidence of superiority.

This study develops a mathematical core that is self-contained. It considers an exact amount finite treatment problem and fixed mean-input periodic examples and a different relaxed exposure distribution bound. Clinical applications and general companion analyses will have their own endpoint and validation contracts and the dimensionless examples are not estimates of patient benefit. Null results, small gains and feasibility restoration are kept because each answers a different scientific question.

## 2 Methods

### 2.1 A matched longitudinal comparison

#### 2.1.1 Dynamics and dose accounting

A regimen s determines a state trajectory x with the modeled PK, benefit, injury, sensitivity and disease components. Parameters are θ. Continuous input u has units of administered amount per unit time; discrete amounts D_i_ enter at times t_i_. Between administrations and across dose jumps, respectively,

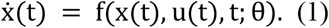

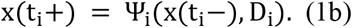

The jump map needs to place a dose in the compartment appropriate to its route. An oral dose normally enters a depot; an intravenous dose can jump a central amount. Absorption and bioavailability belong to the model. Total administered amount is

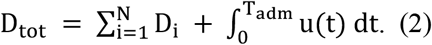

Each dose is counted once. Equal administered amounts do not necessarily produce equal delivered exposure across routes. Infusion amount, rate and duration must satisfy their mass relation, while administration times determine an irregular dosing frequency. The physically admissible class is restricted by the available equipment, dose increments and minimum spacing between administrations.

For well-posed dynamics, suitable regularity, invariance of the physical domain and absence of finite-time blow-up are required. Drug mass should be distinguished from effect-site equilibration or an indirect-response deviation [4,5]. In addition, the observations allow identification only of the states and parameter combinations supported by the measurement model. Practical identifiability is required for predictions used in decision making; agreement with plasma concentration data alone does not establish this property [6].

#### 2.1.2 Objective horizons and path safety

For an efficacy horizon T_eff, integrated and terminal benefit are defined in compatible utility units as follows:

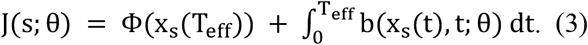

The definition of benefit should specify whether it refers to a direct effect, a reduction in disease burden or another endpoint. When a terminal outcome is combined with its running proxy, appropriate weights are required in order to avoid unintended double counting. Peak injury, cumulative injury, disease outcome, administered amount and treatment burden should be considered separate endpoints, even when they are combined in a scalar objective.

The administration, efficacy and safety horizons are specified separately. Drug input may end while benefit continues, and injury may reach its maximum after both horizons have ended. Therefore, the common safety constraints are applied over the entire prescribed trajectory, including both sides of relevant jumps. A finite penalty alone does not certify compliance with a hard safety constraint. In the case of a mandatory exact amount, reducing that amount does not restore feasibility of the same problem. A resource cap or an objective of minimizing dose defines a different problem.

For finite comparisons, the initial state, total amount and outcome horizon are kept the same. In periodic comparisons, cycle closure is required for every state assumed to be stationary, and cycle averages are calculated at a common mean input. Under linear periodic clearance balance, mean concentration equals bioavailability multiplied by mean input and divided by clearance. This relationship does not generally apply to finite courses or nonlinear clearance. In the relaxed static problem, mean concentration is fixed, whereas kinetic transitions and PD memory are not represented. Thus, the resulting value is an upper bound for an achievable schedule only when the moment constraints and objective are matched.

#### 2.1.3 Incremental value and evidence

The declared reference is included in *A*_0_. Amount choices at fixed times are added in *A*_1_, selected time choices in *A*_2_, and the named input structures and cessation permissions in *A*_3_. Classes must be truly nested. Their common feasibility filter gives

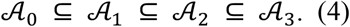

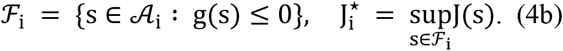

For nonempty feasible sets with finite suprema, inclusion implies nondecreasing optimal values. This result does not establish a strict gain, attainment of an optimum or clinical importance. The class indices refer only to the comparison under consideration. For example, cessation provides no additional choice when terminal zero input is already permitted in the preceding class.

A feasible candidate from an expanded class establishes existence. A strict class gain is established when the candidate objective exceeds a valid upper bound for the smaller feasible class. Global optimization of the extended class is not required for this purpose. When either class optimum is unknown, differences between candidates are reported as observed gains. Failure to identify a feasible solution does not prove infeasibility. Similarly, an infeasible constant strategy does not establish that the entire constant family is infeasible.

Feasibility restoration refers to obtaining a nonempty feasible set while retaining the same target, safety and resource constraints. It should not be confused with an objective difference, which is undefined here if a required feasible set is empty. Demonstrating dose saving requires minimizing the realized administered amount at a common target; a fixed-amount benefit gain cannot establish dose saving. Numerical bounds must take integration and floating-point errors into account, and analytical certificates are only valid for the model and class that we use.

#### 2.1.4 Information uncertainty and computation

A nonanticipating policy makes decisions based on available information. An open-loop schedule belongs to that class of policies only if the action choices allowed by the policy and the initial condition and the operating rules are the same. Feedback value compares the feasible policy and the open-loop optima under one criterion. Information value compares observation filtrations while accounting for measurement costs. Additional information has nonnegative value only if it can be ignored without an unavoidable cost or a change in constraints. Receding-horizon updates are compatible with Bayesian forecasting, but need not solve the full partially observed problem [7,8,12].

Nominal, worst-case optimization and expected-utility optimization answer different questions. A joint chance constraint concerns the probability of any violation across all safety endpoints and the entire horizon; separate pointwise probabilities are not sufficient to provide this assurance. Robust and chance-constrained formulations require certain appropriate uncertainty sets or predictive distributions [17,18]. Their mathematical labels alone do not qualify a clinical model or certify nonlinear trajectory safety.

Independent evaluation of the resulting trajectory must follow optimization. Checks cover jumps, concentration peaks, injury extrema, periodic closure, startup, cessation tails and restart feasibility. Search starts and control parameterizations determine the scope of the numerical evidence. Necessary conditions from optimal control are not sufficient to establish global optimality. Tools such as mapbayr, nlmixr and TUCUXI can provide estimation or prediction infrastructure [10,11,15], while the comparison objective and verification remain separate tasks.

### 2.2 Analytical boundaries and positive benchmarks

#### 2.2.1 Periodic smoothing and a desensitization null

Consider periodic linear PK with a fixed mean input, a directly acting concave benefit b(C) and a convex toxic drive v(C). In this case, the corresponding constant concentration is assumed to be attainable within the input class, with no additional memory, adaptation or time-dependent weighting. Mean concentration is then determined by the clearance balance. With angle brackets indicating cycle averages, Jensen’s inequality gives

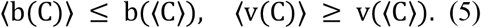

Both bounds can be attained with constant exposure. With linear injury repair, ż = v(C) − ρz and ρ > 0, periodic balance also results in mean injury ⟨z⟩ = ⟨v(C)⟩/ρ. Thus, smoothing provides the highest mean benefit and the lowest mean drive and mean injury under these assumptions. With strict concavity, any genuinely variable exposure gives a strictly lower mean benefit.

This finding refers to the attainable constant concentration in the periodic direct-response system. Finite startup, terminal rewards and models with exposure-dependent repair require separate analysis. The example serves as a negative control: no additional benefit can be obtained by enlarging the class when an existing member already attains the valid bound.

The physical domain [0, 1] is invariant for sensitivity a. Suppose that recovery is independent of exposure, that benefit and desensitization share a bounded, nonnegative concave activation q(C), and that exposure is periodic. Assume k_rec_ > 0, k_des_ > 0 and, on the periodic solution, a(t) > 0 at every time t in the cycle. Since a is strictly positive, division by a and integration of its logarithmic derivative can be performed:

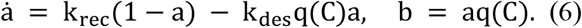

Periodic sensitivity balance and logarithmic balance give an upper bound on mean benefit. Define 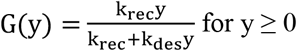. Then

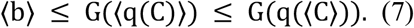

Integrating Equation 6 over one cycle for a positive periodic sensitivity yields ⟨b⟩ = (k_rec_/k_des_)(1−⟨a⟩). Dividing the sensitivity equation by a and integrating its logarithmic derivative gives 0 = k_rec_⟨1/a⟩ − k_rec_ − k_des_ ⟨q⟩. Since the reciprocal is convex, ⟨1/a⟩ ≥ 1/⟨a⟩. Consequently, ⟨a⟩ ≥ k_rec_/(k_rec_+k_des_⟨q⟩), and the benefit identity implies ⟨b⟩ ≤ G(⟨q⟩).

For nonnegative scalar y, G is increasing. Concavity of q therefore implies G(⟨q(C)⟩) ≤ G(q(⟨C⟩)). Constant concentration together with stationary sensitivity attains this bound. The argument applies to the specified periodic benefit model; it does not compare an independent injury process or finite-treatment transients.

Different benefit and desensitization activations, exposure-dependent recovery, finite transients or independent injury constraints require a separate comparison.

#### 2.2.2 A relaxed opportunity from response curvature

For a static Hill benefit, assume h > 1, EC50 > 0, Emax > 0 and a concentration interval [0,C_max_] with C_max_ > 0. The response and its tangent concentration are

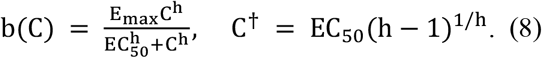

The relaxed problem selects a favorable concentration by maximizing benefit per unit exposure. When the interval contains C†, the concave envelope connects zero to C† and then follows the concave portion of the Hill curve. A mixture of zero and C† attains the envelope for means below C†; constant exposure does so for larger means. If the concentration cap C_max_ lies below C†, the envelope on [0,C_max_] is the straight chord from zero to b(C_max_), and a zero/C_max_ mixture is optimal in the relaxed problem. This follows from the increase of b(C)/C up to C† and applies to means in the given interval. A mean exceeding C_max_ is infeasible.

The mechanism here is exposure allocation, with no recovery state. For the normalized example h = 3, EC50 = Emax = 1 and mean concentration 0.3, the envelope value is 0.158740, compared with 0.026290 under constant exposure. This difference bounds a relaxed opportunity; it does not predict the improvement achievable by a bolus regimen. Elimination prevents instantaneous drug removal, and transitions themselves consume exposure. However, memory, injury and administration restrictions are also crucial factors. These features may prevent a static allocation from being implemented directly as a dosing schedule. For this reason, the envelope is used in LOGOS only as an opportunity bound for matched mean exposure and static benefit. Kinetic realization is addressed in a separate optimization.

The related antimicrobial analysis of Katriel minimizes the exposure under an eradication requirement [16]. Similarity of an ideal concentration structure does not transfer any certificate to the present fixed-mean-exposure objective.

#### 2.2.3 Exposure governed recovery and repair

Recovery can be suppressed during active exposure and low exposure can restore responsiveness or tissue function. For a prescribed exposure trajectory, a useful model has nonnegative rates r, d, v and ρ:

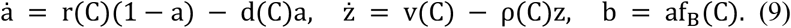

Here f_B denotes nonnegative benefit activation. For a prescribed periodic exposure and locally integrable nonnegative rates, sensitivity remains in [0, 1] and injury remains nonnegative. The integrating factors generate the affine cycle maps a(τ)=A_a a(0)+B_a and z(τ)=A_z z(0)+B_z whose multipliers are exponentials of the negative integrated decay rates and forcing terms are nonnegative. A unique attracting cycle is obtained for each component when its multiplier is less than one. In the case of zero integrated repair and positive injury generation per cycle, injury accumulates and a finite periodic condition cannot be obtained.

An administration pause refers to an interval without new input. However, low active exposure requires clearance of the relevant driver, whereas recovery refers to further improvement in the biological state. These events may occur at different times because of depot input, active metabolites or slow equilibration. A recovery threshold can identify a possible restart time, but does not establish that this time is optimal. The comparison of restart strategies should also account for disease rebound, unobserved responsiveness and predicted injury following the next dose. In addition, periodic closure does not by itself guarantee satisfactory startup or withdrawal.

#### 2.2.4 A globally certified finite Emax benchmark

An ideal model is used as a benchmark in order to assess the broader optimization procedure. Suppose V, k, EC50, Emax, D, T are all positive, volume V is constant, k = CL/V, initial concentration is zero and benefit is Emax C/(EC50 + C). The only input to the model in this case is a nonnegative administered dose measure M of total amount D, which allows for an initial intravenous bolus. The model has no adaptation or terminal payoff and no active rate, concentration or injury constraint. Over efficacy horizon T, concentration satisfies dC = −kC dt + dM/V.

When T exceeds k^−1^log(2 + EC50 V/D), the optimal concentration profile has an initial load to c, a maintenance interval of duration L at c, and an input-free coast of duration δ. Its parameters satisfy

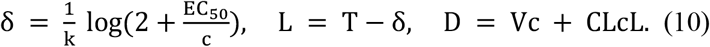

The loading amount is Vc and the maintenance rate is CLc. For a shorter horizon, the optimum is a single initial bolus followed by coast; at the threshold, maintenance duration is zero. The supporting marginal-value inequality below establishes global optimality within this ideal class. During coast, residual drug continues to provide benefit, without any recovery mechanism in the model.

The result follows from a supporting marginal value inequality. We define p(s)=∫_S_^T^ b′(C(t)) exp[−k(t−s)]dt and m(s)=p(s)/V. The adjoint satisfies 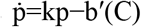, with p(T)=0. On the plateau, m = μ=b′(c)/(kV). The stated δ is obtained by matching the plateau with the input-free coast. During coast, m<μ. Thus, the loading atom and maintained input are supported exactly at times when marginal value is maximal.

For any competing nonnegative dose measure M′ of equal mass, linear PK and concavity lead to J(M′)−J(M) ≤ ∫m(s)d(M′−M)(s) ≤ 0. This inequality provides a sufficient certificate of global optimality within the ideal measure-input class. The same supporting inequality applies to the first atom in the shorter-horizon branch. Due to strict concavity, the optimal concentration trajectory is unique up to sets of zero time measure. A separate solution is required when the problem includes active safety or rate constraints, oral absorption, terminal rewards, or uncertain or nonlinear PK.

During the coast, residual drug continues to provide benefit, although no recovery mechanism is included in the model. Nevertheless, an interval without input that provides benefit need not be favorable when both benefit and harm are considered. This finite-course finding is consistent with the periodic smoothing bound, since the two analyses address different questions.

#### 2.2.5 Safety after cessation

For the cessation tail, assume C_0_, z_0_, α ≥ 0, k > 0 and ρ ≥ 0, with constant parameters. In the one-compartment model, residual exposure is described by C(t) = C_0_exp(−kt), while injury follows ż = αC^2^ − ρz, with no additional input administered or absorbed. Cessation occurs at time zero with injury z_0_. For ρ ≠ 2k, the unique injury trajectory is

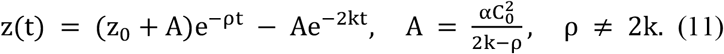

The future maximum is determined by evaluating current injury, all positive stationary times and the limiting endpoint. When ρ=2k, z(t)=(z_0_+αC_0_^2^t)exp(−2kt). In the absence of repair, injury increases toward z_0_+αC_0_^2^/(2k). Neglecting nonnegative repair therefore gives the bound z(t) ≤ z_0_+αC_0_^2^/(2k). These expressions require the absence of further absorbed input; a metabolite or delayed-response model needs its own calculation.

In the normalized case C_0_ = α = k = 1, ρ = 0.2 and z_0_ = 0, the injury at time 0.05 is 0.047340 and below a limit of 0.1. Its final maximum is 0.387132 at time 1.279214. Stopping administration is not enough to make this trajectory safe. In Figure 1 we show the complete decline after the peak and the exact formula justifies the later tail.

**Figure 1.**
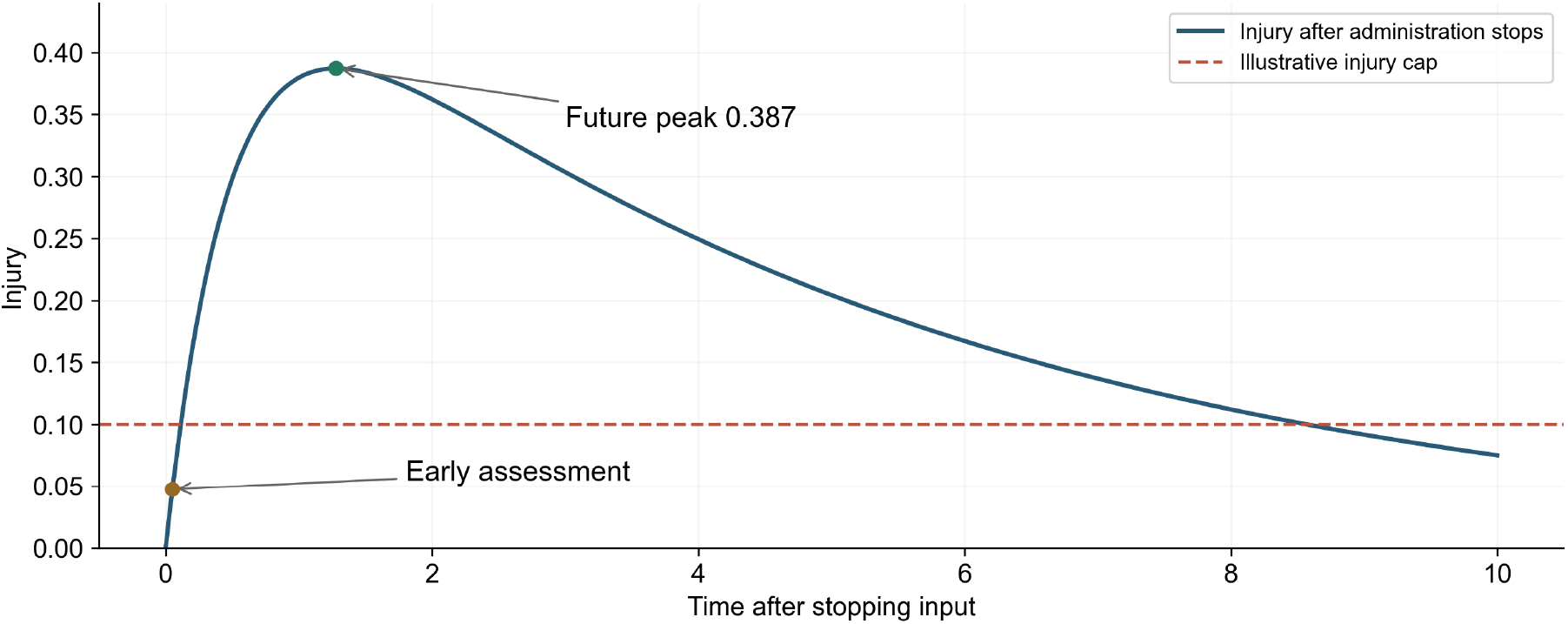
Injury following cessation in the residual exposure model

### 2.3 Numerical examples under explicit comparison contracts

#### 2.3.1 Amounts timing and input structure in a finite course

In this example, V = k = EC50 = Emax = D = 1 and T = 3, with an initial concentration of zero. All schedules are evaluated by cumulative direct Emax benefit on [0,3]. Discrete doses are administered intravenously, with a total amount of one and a concentration cap of one. The injury equation is ż = C^2^ − 0.5z, with an initial value of zero and an upper limit of 0.5 over the entire future tail. Continuous administration is limited to a rate of one. These are nonbinding in this example so we can compare the design freedom rather than the safety benefit. The reference provides three equal boluses at times 0, 1 and 2. In the amount-only class, nonnegative amounts, including zero, are permitted at the specified times. In the timing class, up to three nonnegative boluses are permitted at ordered times within [0, 3]. These schedules are retained in the expanded class, which also permits bounded continuous input. Since terminal zero input is already allowed in the bolus classes, the additional choice in the final comparison is continuous input rather than cessation.

Differences and percentages were calculated from full-precision values; columns are rounded. The verified J values are 0.629606362035524, 0.672945396127757, 0.677043273327298, 0.679552690590195. The last adjacent difference is 0.002509417262897 which rounds to 0.002509. If we subtract already-rounded J columns we get 0.002510.

The fixed-time optimizer allocates around 0.628402 and 0.371598 at times zero and one, and the third amount will be zero. The timing search finds amounts about 0.529424, 0.235288 and 0.235288 at times 0, 0.587748 and 1.175496. Its recorded value is a best candidate from 24 successful multistart runs, not a global timing certificate. The globally certified expanded-regimen benchmark in the above admissible class loads 0.400534, remains at this concentration for 1.496664 and coasts for 1.503336. It is certified by the analytical result.

The adjacent observed gains are 6.8835%, 0.6089% and 0.3706%. Redistributing amounts accounts for most of the improvement over the reference. Relative to the optimized fixed-time solution, the expanded regimen improves benefit by about 0.98%. These small differences represent the tested opportunity and they neither predict a clinical effect nor guarantee increments between unknown class optima. The exposure trajectories and cumulative objectives are presented in Figure 2.

**Figure 2.**
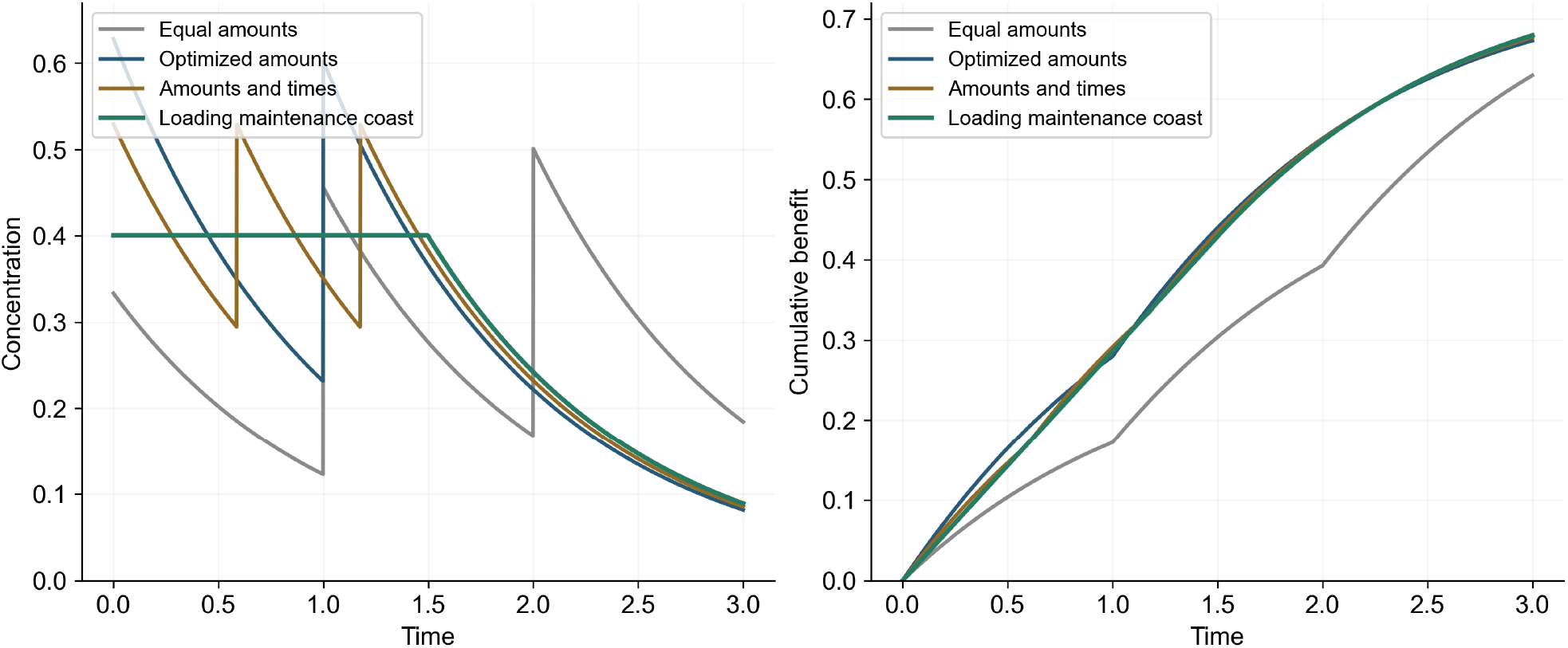
Concentrations and cumulative Emax benefit during finite treatment

For any nonnegative dose measure of mass one with k=V=1, the bounds C(t)≤1 and ∫_0_∞C(t)^2^dt≤1/2 hold. Expanding the squared PK convolution gives us the second inequality: the integral equals one half of the double integral of exp(−|s−r|) against the dose measure twice. Any nonnegative injury repair ensures that z(t) cannot exceed this complete generating-drive integral. Every displayed strategy therefore respects the injury cap over the entire future, without relying on a sampled safety grid. The loading–maintenance–coast candidate also respects the rate cap, since its maintenance rate is 0.400534.

#### 2.3.2 Periodic curvature and joint recovery and repair

For the periodic model, k=V=1 and mean administered input is 0.5. Regular boluses separated by τ deliver an amount 0.5τ and produce a peak concentration of 0.5τ/[1−exp(−τ)]. Benefit activation is q=C/(1+C) or q=C^3^/(1+C^3^), desensitization is d=2q, and benefit is aq. Both cases use the injury drive q_T=C^8^/(0.35^8^+C^8^). Exposure-independent rates are r=0.7 and ρ=1; exposure-dependent rates are r=0.7(1−q)^4^ and ρ=1−q_T. Every case has a concentration cap of four and an injury cap of 3.5.

With concave activation and independent recovery/repair, constant exposure is feasible and achieves the analytical benefit bound; the shortest tested bolus interval approaches it from below. With sigmoidal activation and independent rates, the selected feasible bolus increases mean benefit by 32.22%. With dependent rates, selected bolus witnesses are feasible and stationary constant injury is 17.346653 and violates the cap. Concave activation also allows for this feasibility difference and so we do not require sigmoidality. Recovery and repair are jointly changed and therefore the experiment cannot separately attribute them.

Across 27 local combinations of baseline recovery, repair and injury cap within ±20% of baseline, constant exposure remains infeasible while a tested bolus is feasible. With the cap tightened to one, no feasible bolus is detected in the nine tested recovery/repair pairs. Raising the cap to twenty makes both strategies feasible in six pairs. Not finding a feasible cycle does not prove that all regimens are infeasible. These deterministic parameter sweeps are not an estimated distribution of patient uncertainty.

#### 2.3.3 Computational methods and reproducibility

Calculations were implemented in Python 3.12 using Jupyter notebooks. Finite objectives are evaluated with exact exponential-segment integrals and fixed-time amounts are constrained to sum to one. The amount/timing search uses ordered times, 24 starts, seed 20261003 and an SLSQP tolerance of 10^−13^. A bracketed scalar root solve determines the plateau. The periodic calculations are performed using affine cycle initialization and independent refinement of the peaks. Initially, 121 intervals on [0.08,12] are scanned. Detected feasibility boundaries are then refined and the neighborhoods of candidate solutions are examined. This finite search does not establish a global periodic optimum. The available computational reconstruction specifies Python 3.12, NumPy 2.3.5, SciPy 1.17.1 and Matplotlib 3.10.9. These package versions describe the reconstruction environment and have not been verified as those used in the original analysis. Jupyter Notebook and ipykernel versions are not pinned in the available environment files.

The dimensionless models are specified by the reported equations and parameter values. Agreement between numerical results is used to check implementation, whereas the analytical arguments establish the scope of global-optimality and complete-tail safety claims. Full computational reproduction also requires the original scripts or notebooks, full-precision selected schedules, solver and quadrature tolerances, environment metadata and search results. Exact reconstruction of the selected periodic schedules is limited by the absence of their bolus intervals from Table 2.

**Table 1.**
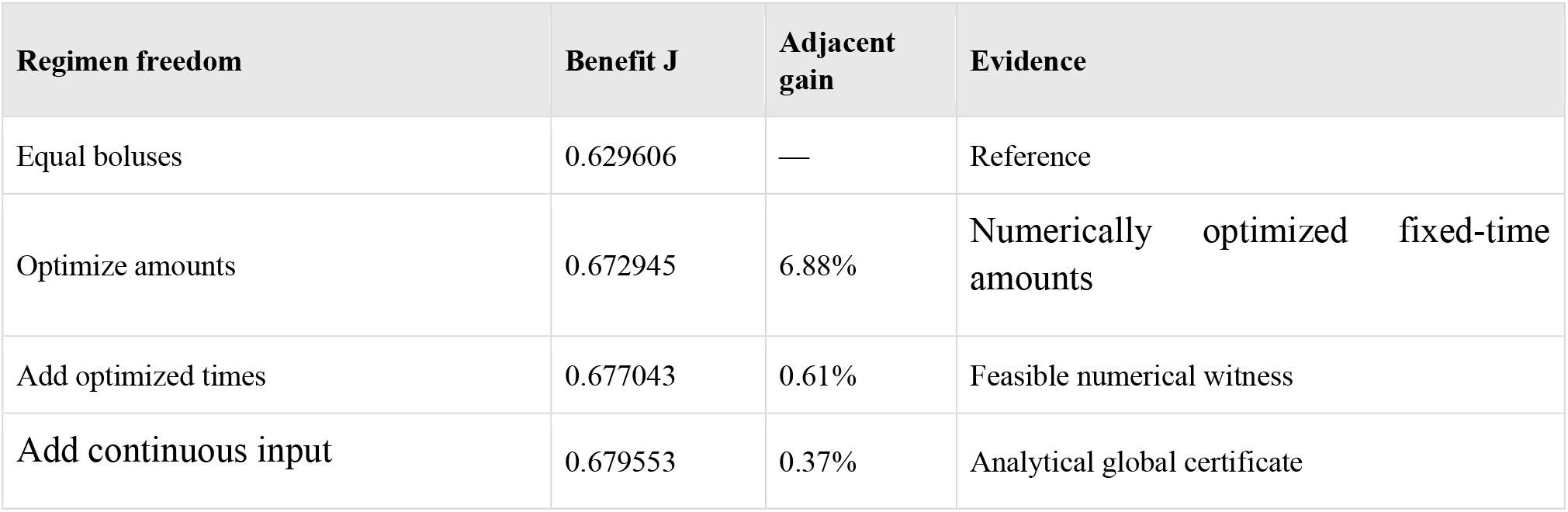
Finite treatment at exact administered amount one.

**Table 2.** Periodic comparisons at matched mean input.

| Case | Constant J | Bolus J | Constant safe | Bolus peak injury |
| --- | --- | --- | --- | --- |
| A concave independent | 0.170732 | 0.170721 | Yes | 0.944709 |
| B sigmoidal independent | 0.084337 | 0.111507 | Yes | 0.766421 |
| C concave dependent | 0.057260 | 0.085510 | No | 1.789967 |
| D sigmoidal dependent | 0.073656 | 0.091278 | No | 1.738049 |

The GitHub repository for the LOGOS general theory provides an online resource for the framework: https://github.com/vkaralis/LOGOS-theory-general.

## 3 Discussion

The aim of the LOGOS approach is to relate an additional regimen decision to an attributable gain, the underlying biological mechanism and the available level of evidence. Its main contribution is the integration of these elements within a matched longitudinal comparison, allowing the source and limits of an improvement to be assessed together. For this purpose, gains due to a larger budget, a different safety limit, a changed horizon or additional monitoring should not be attributed to timing or feedback. A richer parameterization alone does not establish strict value. A simple strategy may already attain the relevant bound, while an improvement between two candidates does not necessarily establish an increment between classes.

The examples presented in this study illustrate different mechanisms. In the specified periodic system, concavity favors smoothing. However, with a finite endpoint, loading followed by an input-free coast can be favored even when benefit is concave. Sigmoidal curvature offers a static opportunity that may not be achievable under kinetic constraints, whereas exposure-dependent repair can alter feasibility. Therefore, these findings should not be considered a general prescription for scheduling. Injury mechanisms should be assessed together with disease rebound, since optimization of recovery alone may favor unrealistic pauses when the consequences of withholding treatment are omitted.

For clinical application, a qualified model for the specific drug is required, together with observations that constrain the relevant states and a comparator representative of actual clinical practice. Counterfactual schedules must also be considered that go beyond the treatment histories used for model fitting. Internal prediction, exposure guardrails, and modeled safety states have their own types of evidence; none can replace evidence of better patient outcomes. Ideal dose atoms and arbitrary continuous inputs also require assessment of whether they can be delivered in practice.

LOGOS complements established optimal control, dose-schedule design and MIPD [1–3,7,13,14]. Its proposed originality is the integrated comparison and attribution protocol, not optimization itself or an assertion that every theorem is new. The combined assessment of null benchmarks, attainable witnesses, global certificates and safety after cessation provides a common basis for evaluating the scientific value of whole-regimen flexibility. In future studies, achievable gains should be presented together with operational burdens, uncertainty and clinically interpretable endpoints. Small or zero increments are also informative, since they identify situations in which additional regimen flexibility is unnecessary.

## 4 Conclusion

The aim of this study was to present an integrated framework for assessing the conditional value of whole-regimen decisions. The LOGOS approach uses matched nested classes, mechanism-specific comparisons and explicit evidence requirements in order to distinguish improved benefit, restored feasibility and absence of added value under the same objective, resources and relevant constraints. The finite benchmark provides a global certificate, while the periodic examples provide model-specific witnesses and null results. In addition, the cessation example demonstrates the need to assess safety beyond the administration period. Thus, the framework allows additional regimen flexibility to be examined through testable comparisons with a clearly defined scope. Its ability to improve prospective patient outcomes remains to be established.

## Data Availability

https://github.com/vkaralis/LOGOS-theory-general

## Ethics approval

Not applicable.

## Informed consent

Not applicable.

## Data availability

The dimensionless examples are specified by the equations and parameter values reported in this paper. No patient observations were studied. The associated Zenodo record provides a persistent identifier for citation and access (DOI: 10.5281/zenodo.23402665): https://doi.org/10.5281/zenodo.23402665.

